# Model uncertainty estimates for deep learning mammographic density prediction using ordinal and classification approaches

**DOI:** 10.1101/2024.08.31.24312184

**Authors:** Steven Squires, Grey Kuling, D. Gareth Evans, Anne L. Martel, Susan M. Astley

## Abstract

**Purpose:** Mammographic density is associated with the risk of developing breast cancer and can be predicted using deep learning methods. Model uncertainty estimates are not produced by standard regression approaches but would be valuable for clinical and research purposes. Our objective is to produce deep learning models with in-built uncertainty estimates without degrading predictive performance.

**Approach:** We analyse data from over 150,000 mammogram images with associated continuous density scores from expert readers in the Predicting Risk Of Cancer At Screening (PROCAS) study. We re-designate the continuous density scores to 100 density classes then train classification and ordinal deep learning models. Distributions and distribution-free methods are applied to extract predictions and uncertainties. A deep learning regression model is trained on the continuous density scores to act as a direct comparison.

**Results:** The root mean squared error (RMSE) between expert assigned density labels and predictions of the standard regression model are 8.42 (8.34-8.51) while the RMSE for the classification and ordinal classification are 8.37 (8.28-8.46) and 8.44 (8.35-8.53) respectively. The average uncertainties produced by the models are higher when the density scores from pairs of expert readers density scores differ more, are higher when different mammogram views of the same views are more variable and when two separately trained models show higher variation.

**Conclusions:** Using either a classification or ordinal approach we can produce model uncertainty estimates without loss of predictive performance.

## 1. Introduction

Mammographic density is a strong independent risk factor for breast cancer [1] and improves predictive performance of risk prediction models [2]. There are many approaches to density measurement, some of which are categoric, for example the subjective Breast Imaging Reporting and Data System (BI-RADS) [3]. A continuous measurement based on the average of a pair of expert medical practitioners’ subjective estimates of percentage density as recorded on a Visual Analogue Scale (VAS) has been shown to have a stronger relationship with the risk of developing cancer than other methods evaluated [4]. Superior performance at cancer risk estimation is desirable but VAS scoring is not scalable for screening applications due to workload and level of expertise required.

Automated methods would enable VAS-like measures to be more widely used and may also provide more consistent scores [5]. Deep learning models have been trained that show good correlation with expert reader VAS scores along with comparable breast cancer risk prediction performance [6, 7]. These studies demonstrate the potential for deep learning models to be used in clinical practice. However, these methods lack an estimate of model uncertainty in prediction. This is a feature of standard regression tasks where a point output is defined as the target.

For clinical practice model uncertainty estimates have a number of potential applications. One example is for the model to flag any image with a relatively high uncertainty estimate to enable an expert reader to visually assess the density estimate. Another application might be to utilise both the prediction and uncertainty to enable clinicians to review the assignment of women to risk categories. For example, a woman in a low-risk category but with a high model uncertainty estimate might be reassessed due to the possibility of the model having underestimated her risk. The uncertainty of a risk estimate could also be communicated with both medical personnel and the patient which could aid with decision making [8].

Model uncertainty estimates also have value for further research. One potential training approach would be to place more emphasis on images that had shown high uncertainty in previous trained models, either through weighting the images in the objective function or by over-(or under-) sampling images. The uncertainty estimates could also be directly included as an additional term in the objective function to force the model to increase its confidence in its predictions.

Each label in the dataset is produced by averaging a pair of experts’ scores and there is known to be substantial reader variation [9]. The effect of the variability on deep models can be substantial [5] and uncertainty estimates may provide guidance on how to deal with these effects. It may be possible to use known reader variability alongside model uncertainty to separate out uncertainty caused by the reader variability from other types of uncertainty. Potential methods for correcting variability [10] could also be tested for their effect on reducing model uncertainty. Low dose mammograms can also be used for density prediction [11, 12] and uncertainty estimates might be particularly useful due to increased levels of noise in the images which may increase prediction variability.

Previous work has investigated uncertainty and reliability [13] in mammographic density prediction using deep learning. This work investigated how model predictions differed from expert reader estimates considering certain factors, such as the amount of breast in the image. This type of approach provides guidance on average prediction reliability for different factors. For example, they showed that model predictions on denser and smaller breasts were less reliable (showed greater variation compared to expert reader scores). However, these reliability estimates are averages and not specific to individual images and women. The model uncertainties we provide are for individual images.

Or aim is to produce estimates of model prediction confidence on individual images without reducing model predictive quality compared to a standard regression method. Our approach is to consider the problem as categoric rather than continuous. We develop three deep learning models with different labels and objective functions: one as a standard regression and two via a binned classification approach where one of the two is trained as a standard classifier and the other as an ordinal classifier. We both apply a distribution to the data and use a distribution-free approach on the two classification approaches which enable both predictions and model uncertainty to be estimated.

## 2. Methods

The standard regression model is trained using mean squared error (MSE), throughout the paper we refer to this as the *regression* model. The first model that produces uncertainty estimates uses VAS scores separated into 100 bins as labels with the model trained as a classification task, which we refer to as the *classification* model. The last model is an ordinal model where we train on the same classified bins but in a hierarchical manner, referred to as the *ordinal* model. We first discuss those aspects that are common to all three models then describe the characteristics of the individual models. As far as possible, many aspects of the model and training are kept the same so that the comparisons are fair.

### 2.1. Data

We use data from the Predicting Risk of Cancer At Screening (PROCAS) [14] study considering 151,806 mammographam images. In PROCAS, two expert readers (radiologists, advanced practitioner radiographers and breast physicians), drawn from a pool of 19, provided percentage density scores recorded on 10cm Visual Analogue Scales (VAS) for each image and the VAS score per woman was produced by averaging scores for all mammographic projections and the pair of readers. For an individual mammographic image, the pair of reader estimates for that view were averaged.

We partition the data with 101,316 images for training, 25,332 for validation and 25,158 for testing. The images from one woman are kept together so that the same women does not have separate images in different partitions.

There are two different image formats in the PROCAS dataset, all from GE Senographe Essential machines: 2294 *×* 1914 and 3062 *×* 2394. These are padded and cropped into the same size, then reduced to 640 × 512 by cubic interpolation, with image intensities clipped to 75% of maximum and the pixel intensity inverted before histogram equalisation is performed and the pixel intensities are set between 0 and 1. The images are also normalised before being run through the model (see Section 2.2).

### 2.2. Base model

For all three models we use the ResNet-18 [15] model as a base. ResNet is a popular family of models that have shown high performance across multiple tasks. The specific choice of ResNet-18, as opposed to larger networks like ResNet-50 or ResNet-121 is motivated by choosing a model that has sufficient capacity but that we have enough data to effectively train. In addition, it is argued that smaller, bespoke networks can be preferable (or at least comparable) to these larger pretrained networks [16]. Our choice trades off the competing arguments of larger models versus smaller. The training time and computational requirements are also reduced compared to larger models.

The model weights are initialised from those previously trained on ImageNet [17]. Good correlation between model predictions and expert reader VAS scores using ImageNet trained features alone with linear regression has been shown [7]. The model representation produced by training on ImageNet without fine tuning are capable of producing good mammographic density prediction.

The final fully connected layer is removed and replaced with either one neuron (for the regression model) or 100 neurons (for both the classification and ordinal models). An individual image (see Section 2.1 for details) is copied across the three channels and normalised to have means of (0.485, 0.456, 0.406) and standard deviations of (0.229, 0.224, 0.225) to match the ImageNet images the models were pretrained on.

### 2.3. Training

The training method discussed here is the same for the three models. We use the Adam optimizer [18] and the model weights are saved every epoch if the validation error (defined as the objective function for that model) is lower than any previous value. Model selection is performed by comparing the mean squared error (MSE) between predictions and VAS labels on the validation data. For the classification and ordinal models this is after the final prediction estimates are made (see Section 2.5 and 2.6). We train all our models using a NVIDIA V100-SXM2-16GB GPU.

While data augmentation is an essential tool, different researchers utilise various methods [19] with little consensus as to the best approach. A popular option, applying rotations, is non-trivial due to the nature of our images. Other options such as addition of noise generally may not be effective [20]. Therefore, when training we perform data-augmentation via up-down and left-right flips both with 50% probability.

### 2.4. Regression model

The regression model is designed to act as a direct comparison to the classification and ordinal models. We remove the final fully connected layer which, in the standard ResNet, maps from the 512 dimensional feature vector to 1,000 dimensional classes and replace it with one output neuron with no additional function applied. We then train with MSE as our objective function.

When performing inference the model outputs a value, 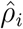, for the *i*^*th*^ image, which we take to be the density estimate. There is no limit placed on the output to force it between 1 and 100 so the results could fall outside of this range but in our experiments this is a rare occurrence.

### 2.5. Classification model

For the classification model we convert the VAS scores per image into 100 classes each representing one of the density scores. Therefore the input data is a set of 100-dimensional vectors with one index taking the value 1 and everything else 0. The model produces the same sized (100-dimensional) output prediction and we apply a softmax to the output and train our model using cross-entropy loss.

This approach factors in no additional information about the relationship between the positive bin and its surroundings. The loss will give the same quantity of error to probability density being placed into a bin next to the labelled bin as a bin further away. Alternative approaches would require the imposition of a prior on the probability distribution we will be learning, as we would need to define how much we would additionally penalise the model for predictions further away from the correct bin. The ordinal model we consider will factor in additional penalisation for distance from the label.

The ResNet-18 model is adapted similarly to the regression model. The final fully connected layer is removed and replaced with a set of 100-neurons each representing one of the VAS classes.

When performing inference we need to make estimates of both the prediction and the uncertainty level of the prediction. We do this both by applying a distribution and in a distribution-free manner.

For the distribution-free estimate we can make predictions using the expectation:

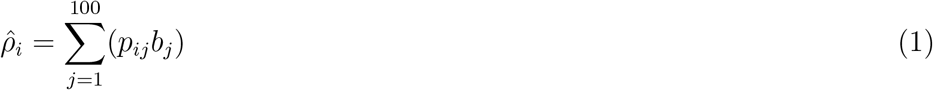

where 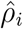 is the model prediction for the *i*^*th*^ image, *p*_*ij*_ is the probability estimate of the model for the *j*^*th*^ bin and *i*^*th*^ image. *b*_*j*_ is the VAS value associated with each of the 100 bins (values 1 to 100). Another option is to take the bin with the highest probability density but this tends to produce more variable predictions as it overemphasises the random nature of small changes in the probability weightings. All results reported in this paper are from the expectation.

To make our estimate of the uncertainty we consider the standard deviation:

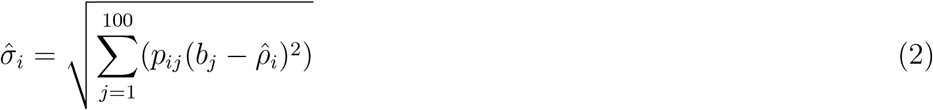

where 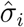 is the uncertainty estimate for image *i*. We consider one standard deviation as the uncertainty but fractions or multiples of that could be chosen and the choice may depend on the required confidence required in the results. We are exploring the general ability of these models to produce uncertainty estimates rather than a more specific requirement of some set uncertainty level.

We also apply a probability distribution to the prediction outputs. We fit the distribution to the data using a maximum likelihood estimate. There are multiple distributions that could be fitted but for simplicity we utilise the gamma distribution which has a probability density function, *f*, of:

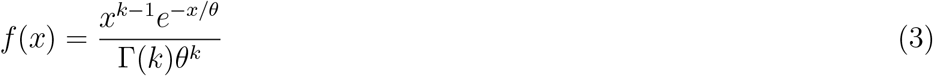

where *k* and *θ* are parameters to be fit and Γ is the gamma function.

The predictions are bounded at both ends, while the gamma distribution is unbounded at one end, so it is not strictly correct to use this distribution but it can fall off fast in its tail so it should be able to do a reasonable job of modelling the data. As the gamma distribution is not symmetric we apply it once to the prediction output without alteration. Then we flip the prediction by applying: 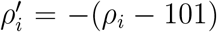 and then fit the gamma distribution again to 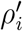.To choose the best distribution we perform the Kolmogorov-Smirnov (K-S) test and choose the one that produces the highest K-S statistic. We do not consider the actual quality of the fit, merely which of the two versions fits better. We will see that the differences between the distribution and distribution-free results are small.

Once the parametric model is selected we then can extract a mean, given by 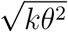, and a standard deviation, given by *kθ*^2^, from the distribution to use as our output prediction and uncertainty, respectively. If the flipped distribution is used then the prediction is flipped back to provide the final prediction.

### 2.6. Ordinal model

The ordinal model approach is based on previous work applying neural networks to ordinal regression [21]. The label for each image is a 100-dimensional vector but instead of having 1 non-zero element all elements up to and including the labelled index are marked as 1. As an example for a 3-dimensional vector consisting of values 1, 2, 3 and a specific sample with value 2, the change from the classification label to the ordinal label would be

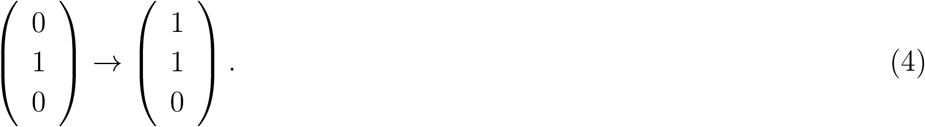

The model produces a 100-dimensional output, in a similar way to the classification model, but a sigmoid is applied to each output neuron independently. For this model the order of the neurons is important. We train the model using the MSE between the label and model output vectors.

When performing inference the distribution-free prediction is made by scanning across the output vector and stopping at the point that the probability falls below 0.5. An alternative is to take the furthest point along the vector that is above 0.5, which could, in principle, produce significantly different results. This could occur if the earliest (along the vector) element that was above 0.5 was some form of outlier or some fluctuation. It could also occur if the predictions do not follow a fairly monotonic reduction as we scan along the output vector. In practice neither of these issues appear to be a problem. We therefore report the prediction as the label of the bin before the first element falls below 0.5.

The uncertainty is estimated by finding the locations of the bins where the desired upper and lower probability bounds are located. We use the 68% (the standard deviation of a Gaussian) confidence interval. The bins where the upper bound first goes below 68% and the lower bound goes below 32% are selected. The confidence interval is the range of the values of the two selected bins.

For the parametric fit we utilise a logistic function. The ordinal model produces predictions that look like a flipped logistic curve with values close to 1 falling to values close to zero. We therefore flip each value such that 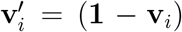 where **v**_*i*_ is the output of the model for the *i*^*th*^ image and **1** is a vector of ones the same size as **v**_*i*_ (100-dimensions). We then apply a logistic function: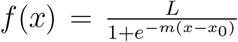 where *L, m* and *x*_0_ are parameters to be found.

The parametric prediction is given by the input value where the probability first falls below 0.5. The uncertainty is defined as the standard deviation of the density of the logistic distribution 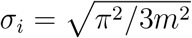. For the model results presented a small number of the predictions (21 out of 25,158) could not be fitted with a logistic curve. For those values the predictions and uncertainty are substituted with the distribution-free values.

### 2.7. Assessment of model performance

To assess the quality of the model predictions we consider the RMSE and Spearman rank correlation coefficients between the model predictions and the expert reader VAS scores. However, the overall metrics may conceal differences at different density scores. To test this we bin the images using a sliding window (of stride 1) of 10 VAS points. The RMSE between model predictions and VAS scores are calculated for the images in the bin. All sample confidence intervals are found via bootstrapping and shown at the 95% level One approach to assess the quality of the uncertainty estimates is to compare them with the differences between the predictions and the expert assigned VAS scores. To do so the data is split up into six bins using the model uncertainties. Within each bin the absolute differences between the predictions and VAS scores are calculated. If the model predictions have a higher uncertainty we would expect there to be a larger average difference between predictions and the VAS scores. We also do the same analysis but using the model predictions from the regression model rather than the expert reader VAS scores.

A further comparison for the uncertainties is how the uncertainty estimates compare to differences in model prediction between the four mammographic views (right craniocaudal, left craniocaudal, right mediolateral oblique and left mediolateral oblique). This is performed for each individual image. We find the average density prediction of the other views and calculate the mean absolute difference to the image prediction. The argument here is that, generally, the four views should show similar prediction to one another. If there are differences between the mammographic views it may imply model variability and therefore a larger uncertainty.

## 3. Results

In Section 3.1 we analyse the predictive quality of the three approaches on the test set. In Section 3.2 we show results for the model uncertainty scores and make comparisons to other factors that may affect uncertainty.

### 3.1 Quality of the models

In Table 1 we show metrics for the final model predictions against labels (expert reader VAS scores). The classification and ordinal approaches show no reduction in quality compared to the regression method. In Figure A1 in the appendix we show plots of model predictions against labels for these results.

**Table 1.** Root mean squared error (RMSE) and Spearman rank correlation coefficient (*ρ*) of the models compared to the VAS score labels. The sample uncertainties are found via bootstrapping and shown at the 95% level.

| | RMSE | $\rho$ |
| --- | --- | --- |
| Regression | 8.42 (8.34-8.51) | 0.843 (0.839-0.847) |
| Classification - distribution-free | 8.38 (8.29-8.46) | 0.845 (0.840-0.849) |
| Classification - distribution | 8.37 (8.28-8.46) | 0.845 (0.840-0.849) |
| Ordinal - distribution-free | 8.38 (8.29-8.47) | 0.846 (0.842-0.850) |
| Ordinal - distribution | 8.44 (8.35-8.53) | 0.846 (0.842-0.850) |

The differences between predictions and labels at different density scores are shown in the left plot of Figure 1. The regression, classification and ordinal models are represented by *R, C* and *O* respectively and the distribution-free and distribution approaches by NP (non-parametric) and P (parametric) respectively. The pattern is the same for all the models with reducing apparent model performance (increasing RMSE) as the density value rises but with improved model performance (reducing RMSE) at the highest density scores. In the right plot of Figure 1 we show the number of images at each expert-assigned VAS score to demonstrate the small amount of data at the higher density values.

**Figure 1.**
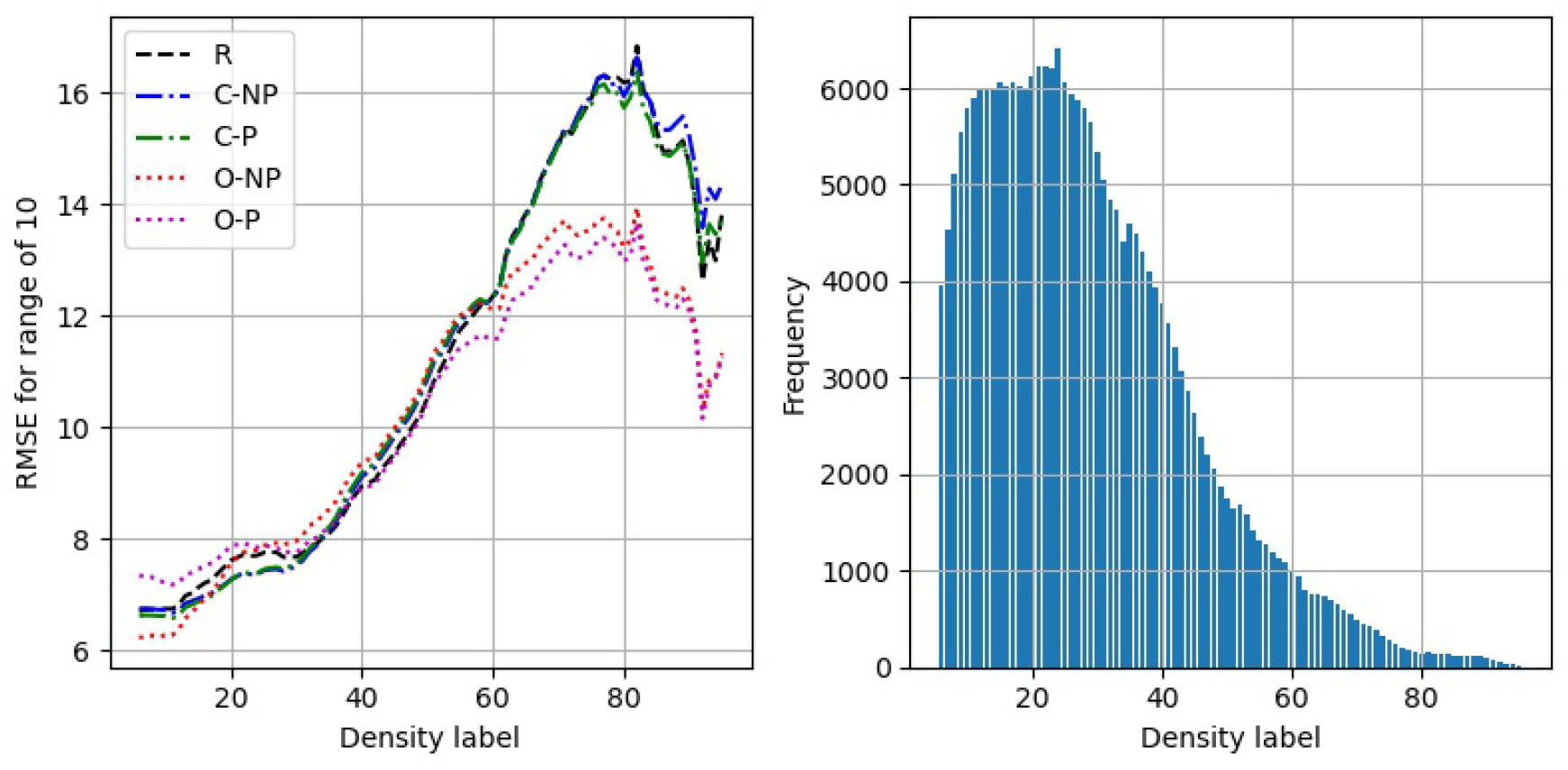
Left) RMSE between predictions and labels at different expert assigned VAS values. Regression, classification and ordinal models are represented by *R, C* and *O* respectively. Right) Distribution of the number of images at each expert-assigned VAS value.

We show the RMSE and Spearman rank correlation between the five sets of predictions in Table 2. The distribution to distribution-free results for the same model are similar especially for the classification model. There is also considerable similarity between all the different sets of predictions. Plots of different model predictions against one another are shown in the appendix (Figure A2).

**Table 2.**
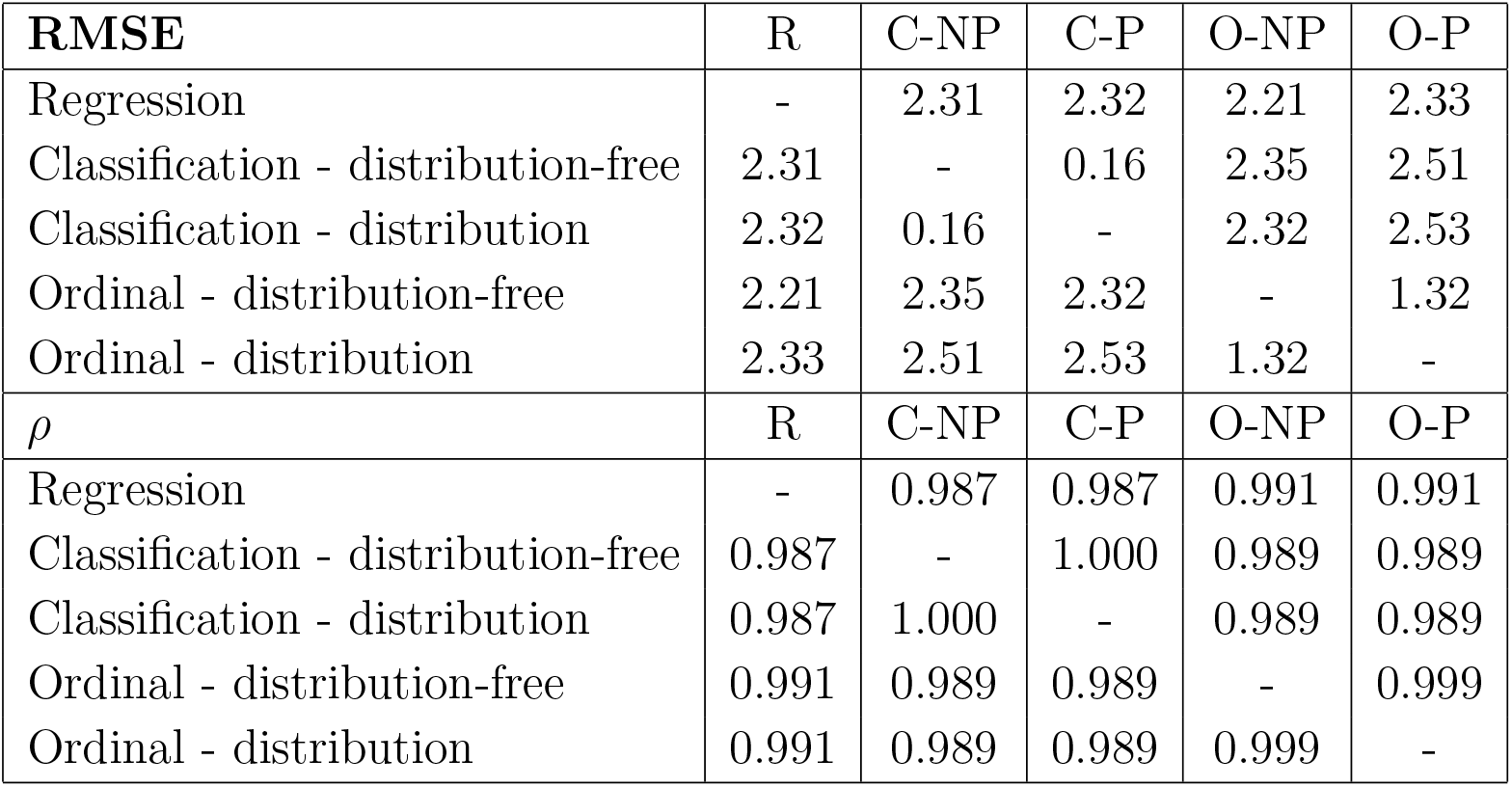
RMSE and Spearman rank correlation coefficients (*ρ*) between the different sets of predictions. All models produce similar predictions.

### 3.2. Uncertainty estimates

There are no uncertainty estimates for the regression approach so we only consider the classification and ordinal models. In Figure 2 we show the uncertainty estimates for distribution (*P*) and distribution-free (*NP*) approaches of the classification model. The left plot shows a histogram of the uncertainties, there is a similar distribution of uncertainty with generally slightly higher values for the non-parametric model. The centre and right plots show direct plots of the uncertainty against the density prediction. The trend is the same for both with rising and then falling uncertainty estimates with prediction value.

**Figure 2.**
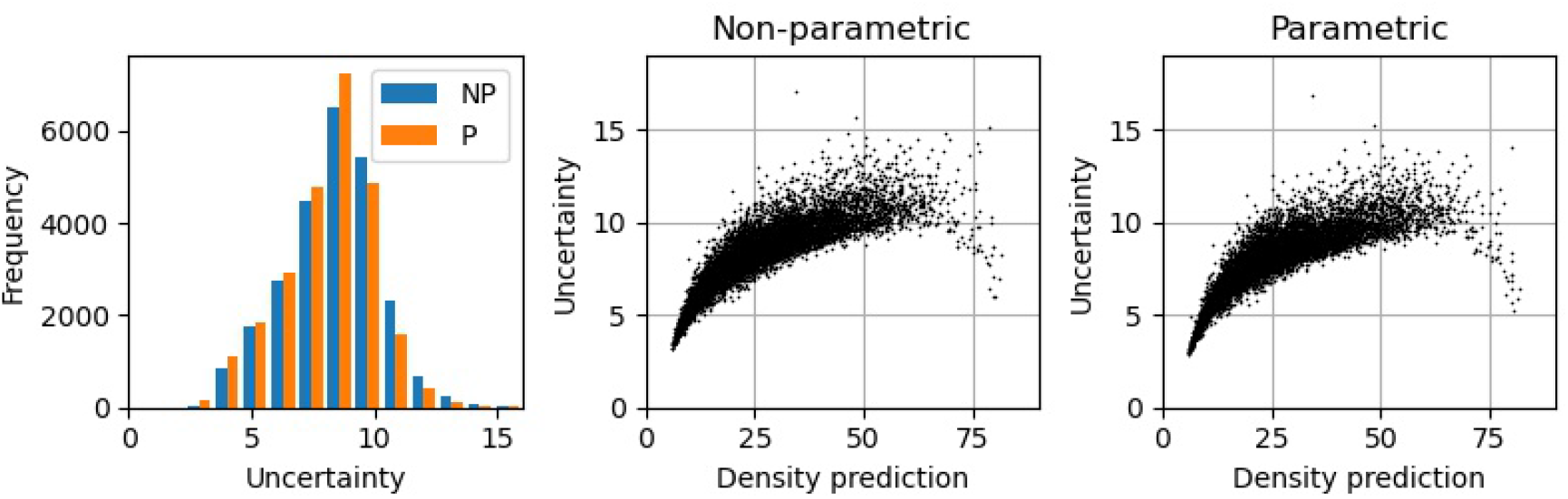
Uncertainty for the classification model. Left) the distribution of the uncertainty values, NP is non-parametric and P is parametric. Middle and right) plots of uncertainty against the predictions for non-parametric and parametric versions respectively.

In Figure 3 we show uncertainty versus density prediction for the ordinal model with both parametric and non-parametric results shown. These are the equivalent results for the ordinal model as Figure 2 was for the classification model. The differences between the non-parametric and parametric models are more pronounced than for the classification model.

**Figure 3.**
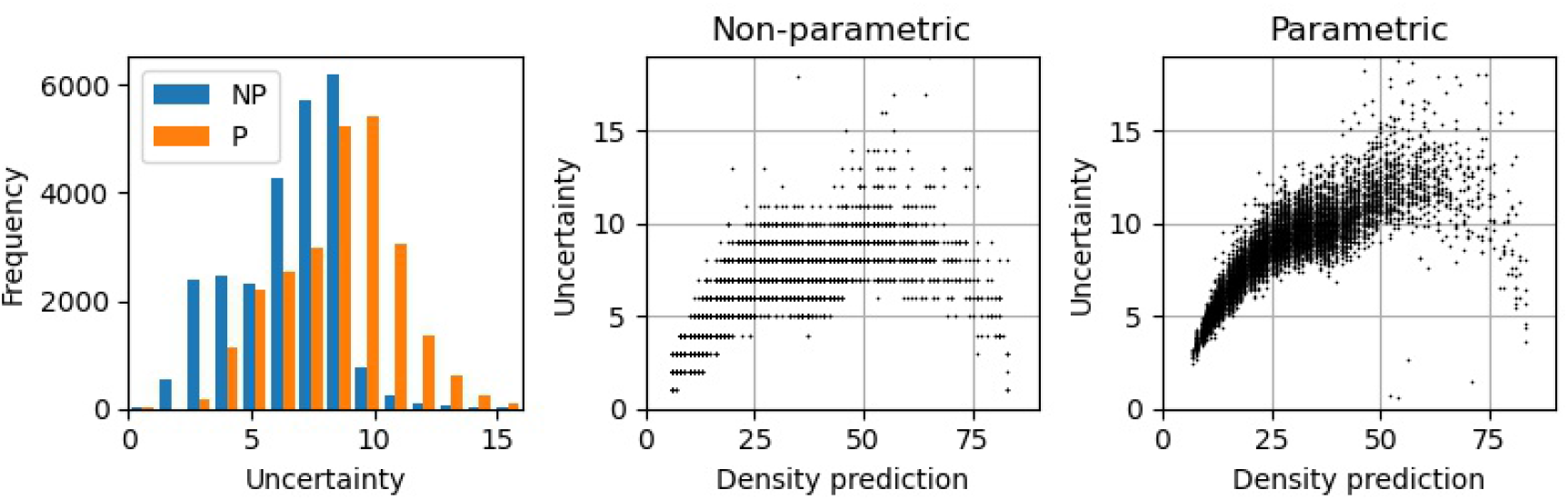
Uncertainty for the ordinal model. Left) the distribution of the uncertainty values, NP is non-parametric and P is parametric. Middle and right) plots of uncertainty against the predictions for non-parametric and parametric versions respectively.

To explore these differences further, in Figure 4 we show uncertainties versus uncertainties for the different models. The classification pair of models (parametric and non-parametric) produce almost identical uncertainty estimates with a small number of points away from the trend-line. The trends for all comparisons are similar - if one uncertainty estimate is high we would expect the other to also be high.

**Figure 4.**
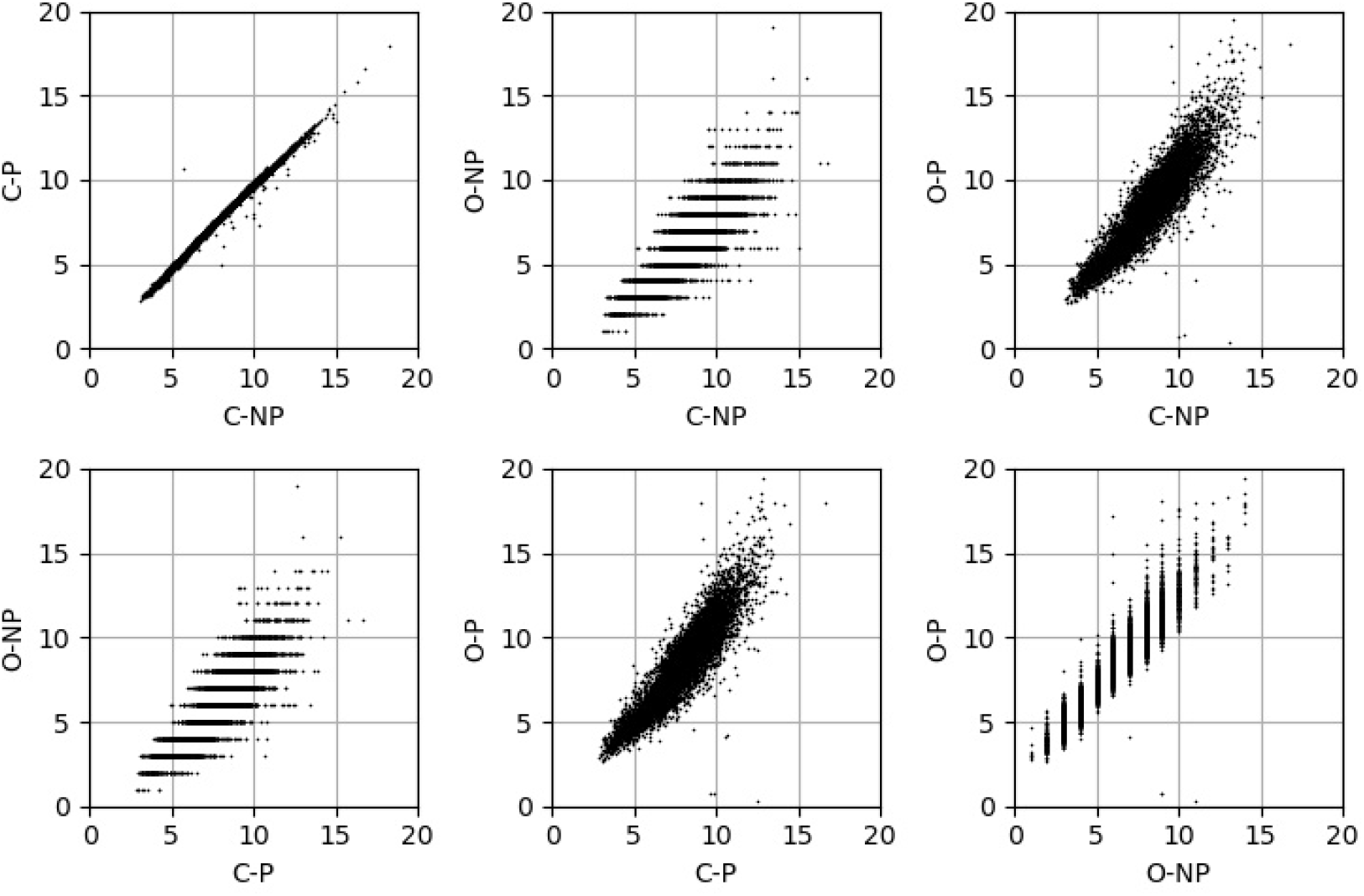
The model uncertainties plotted against one another. C is classification method, O is the ordinal method. NP is the non-parametric method and P the parametric.

The differences between the predictions and the labels for binned model uncertainty estimates are shown in Figure 5. Results for the classification model are shown as the top plots and the ordinal model on the bottom. The left and middle plots show boxplots of the absolute differences for the distribution-free and distribution approaches respectively. The right plot shows the average of the absolute differences for each bin. There is an increase in the average of the differences between labels and predictions as the model uncertainty increases. The classification (top row) and ordinal models (bottom row) show similar results. We show the equivalent plots but with regression model predictions replacing the labels in Figure A3 of the appendix with a similar pattern.

**Figure 5.**
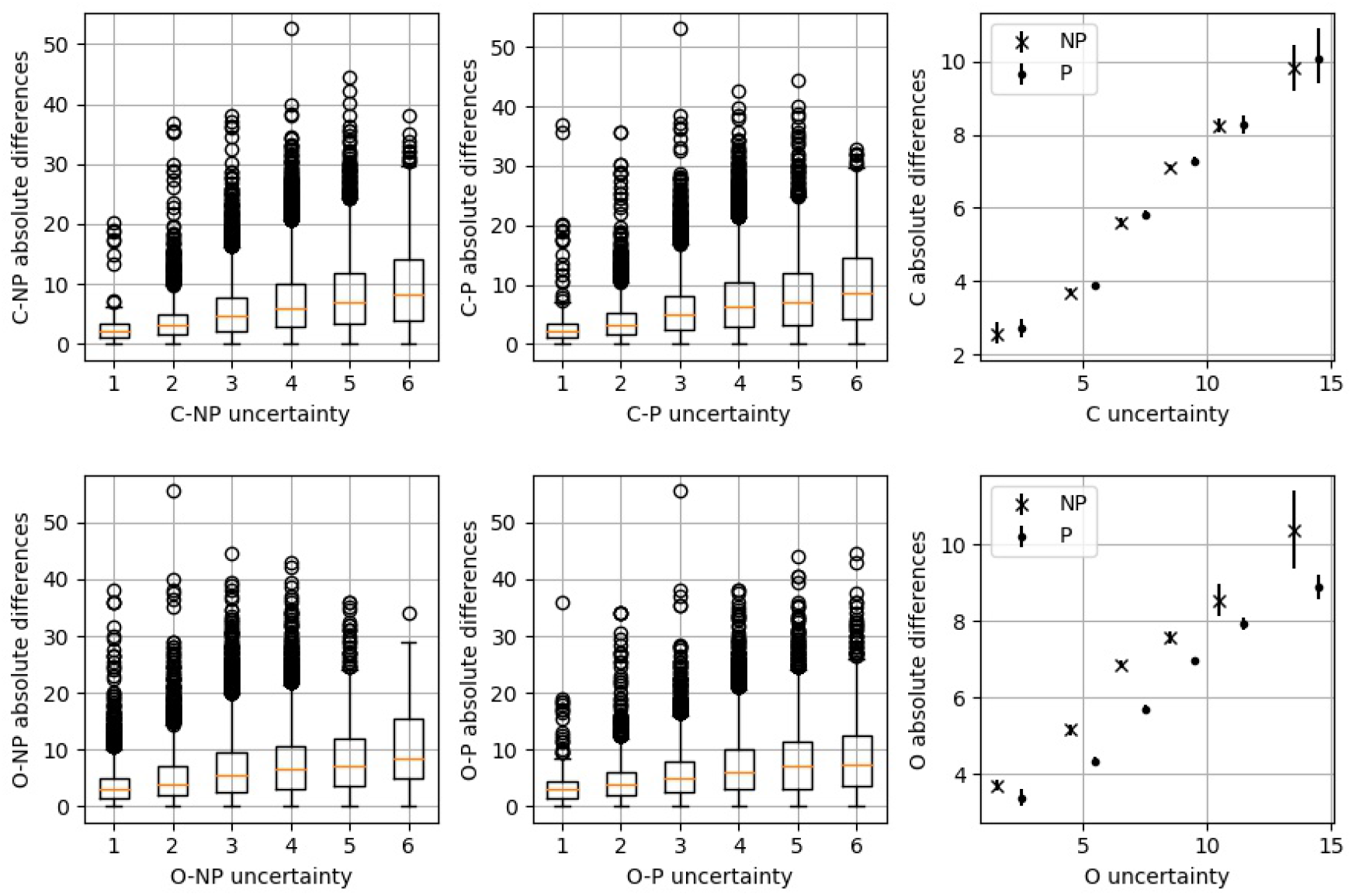
The model uncertainties are split into six bins and the absolute differences between labels and predictions are shown. Left and middle) boxplots showing the absolute differences for the different uncertainty binned results. Right) averages of the absolute differences between predictions and labels in each of the uncertainty bins. The sample uncertainty bounds are produced via bootstrapping at the 95% level. Top plots are for the classification model (C) and bottom plots for the ordinal model (O). NP is non-parametric and P is parametric

In Figure 6 we show uncertainty estimates compared to differences in model prediction between the four (RCC, RMLO, LCC, LMLO) views. These results are in the same format as Figure 5.

**Figure 6.**
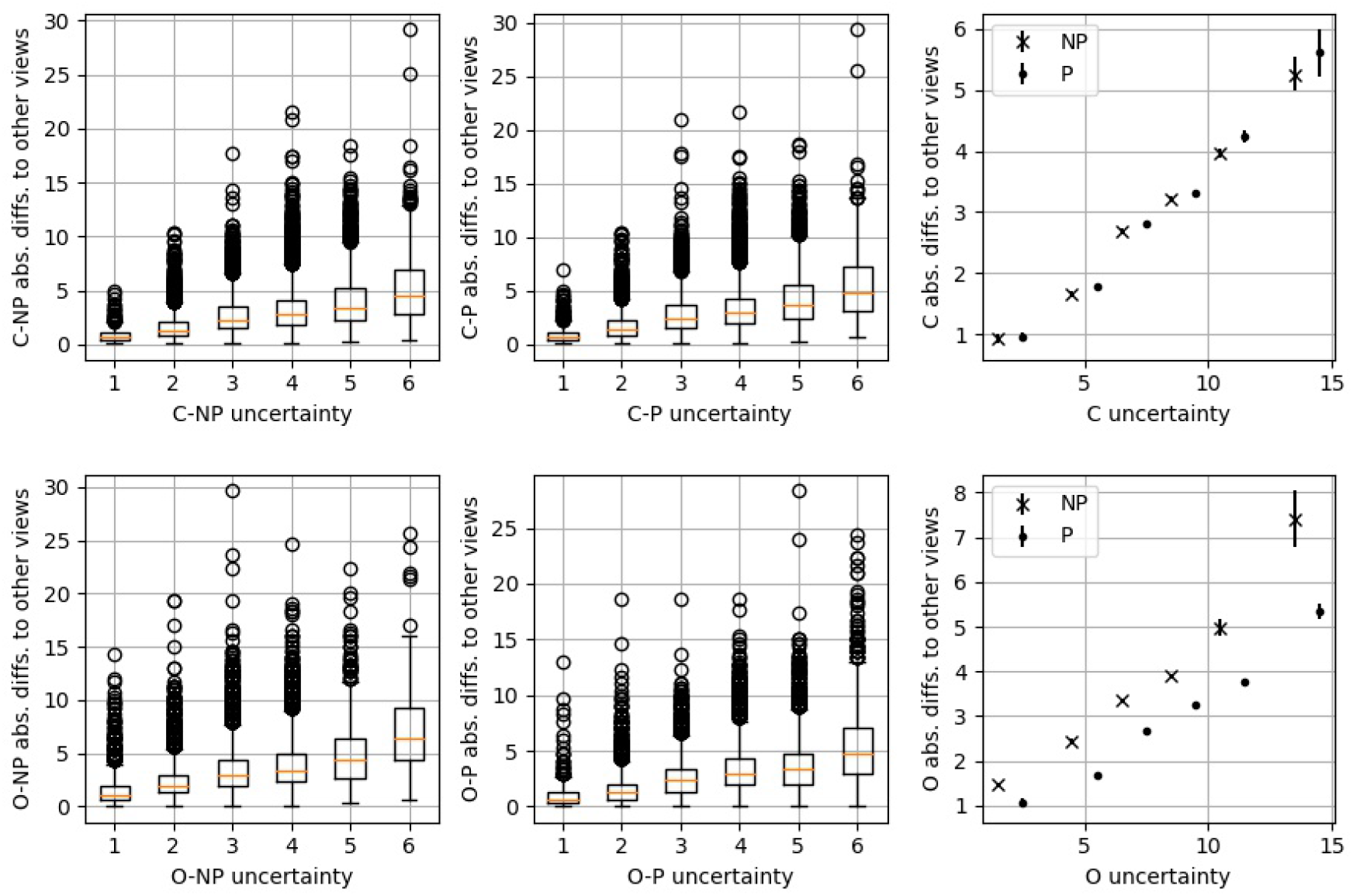
The model uncertainties are split into six bins and the absolute differences between the predictions made on one view and the average of the other views are calculated. Left and middle) boxplots showing the absolute differences for the different uncertainty binned results. Right) averages of the absolute differences between one image and the average of the other views of the same woman in each of the uncertainty bins. Top plots are for the classification model (C) and bottom plots for the ordinal model (O).

## 4. Discussion

We demonstrate two model approaches to enable uncertainty estimation: a classification model and an ordinal model. In addition, we used both non-parametric (distribution-free) and parametric (with a distribution applied) methods to extract the predictions and the uncertainties. We also compared these methods to a standard regression approach with a mean squared error objective function. The inclusion of this regression model enables us to assess whether the classification or ordinal models are losing predictive performance compared to a standard regression approach.

The prediction quality produced by the classification and ordinal model are similar to those of the regression model for both non-parametric and parametric versions. No prediction quality is lost by moving from a regression model to a classification or ordinal approach. There are differences in performance between the models at different densities with slightly improved performance at the higher density range for the ordinal model. These results would have little effect on the overall metrics due to the small number of images at high densities.

The classification model makes predictions of similar quality as the regression or ordinal approaches. This is despite the probability density of the neurons in proximity to the positively labelled neuron providing feedback that the predictions are equally as wrong as those neurons far away. While a clear explanation of why the classification model is able to make such accurate predictions is beyond the scope of this work we do suggest the density signal might be large compared to anything else in the image. Previous work has demonstrated that deep learning models can extract the density signal even if the labels are variable [5]. These classification results imply even with a small positive signal the model can learn a good representation.

We showed that the uncertainty for all the models correlates with differences to labels, differences to the standard regression model and also to the differences to other views from the same woman. The results all show the pattern we would expect if the uncertainties were correctly estimating a true uncertainty. In particular the uncertainties being correlated with differences between predictions and labels is significant because when using these models we would usually not have available expert reader VAS labels. The distribution of uncertainty across the density prediction distribution (see Figure 2 and Figure 3) show a similar distribution to the differences between pairs of expert reader density scores [6, 7]. This may imply that some of the increased uncertainty is being caused by the reader variability or that some other issue with mammograms causing both expert readers and the models to have greater uncertainty. The distributions of uncertainty across the density prediction range may also be partially a feature of the production of the prediction estimates, especially for the classification approaches. For the non-parametric version of the classification method the expectation is used to make the prediction. To produce predictions close to either end of the density distribution requires there to be limited probability density placed in bins far away from the expectation otherwise the expectation would be shifted away from the ends of the distribution. This means that the uncertainty cannot be too high for the images at the ends of the distributions.

Our results show little evidence to enable us to choose between the two uncertainty models or between the parametric and non-parametric approaches. The quality of prediction of the classification and ordinal methods are similar and show no statistically significant differences. We can conclude that both methods appear to produce uncertainty estimates that show the sorts of behaviour we would expect to see. When considering the two approaches the ordinal method seems intuitively like it should produce better results due to the issues that we have discussed for the classification method. However, we do see any evidence in the data. Therefore we present both methods as equally valid for uncertainty estimation.

In the introduction we specified multiple potential uses of uncertainty estimates for mammographic density predictions with brief descriptions of the approach to take. We have demonstrated the capacity of these approaches to produce model uncertainty estimates with no loss of predictive power. The detailed specifics of how to utilise the uncertainty estimates for clinical uses and to further improve the models will need to be further investigated.

## Data Availability

Data is not currently available to researchers outside the University of Manchester.

## Appendix A. Additional results

Plots of the predictions versus labels in Figure A1 with *NP* and *P* standing for the non-parametric and parametric models respectively. We show a random selection of 5,000 out of the 25,158 test images for ease of viewing. Similarly to the results in Table 1 there is no evidence of reduction in performance when using either the classification or ordinal models compared to the standard regression.

In addition to the metrics in Table 2 we also show, in Figure A2 how the five model predictions compare to one another, as with Figure A1 we show just 5000 points. The are some systematic differences, for example between the regression and ordinal models. The different models also show some noise between them but the general trends of all are similar.

An additional comparison we make to try and elucidate the quality of the uncertainty estimates is to compare to the predictions made by the regression model. We perform the same approach as to the labels but substitute the regression prediction. The results for these are shown in Figure A3 with all the same plots as in Figure 5. We see a very similar pattern - where the uncertainties are higher we see larger differences in prediction to the regression model - note that the y-axis is differently scaled between Figures A3 and 5.

**Figure A1.**
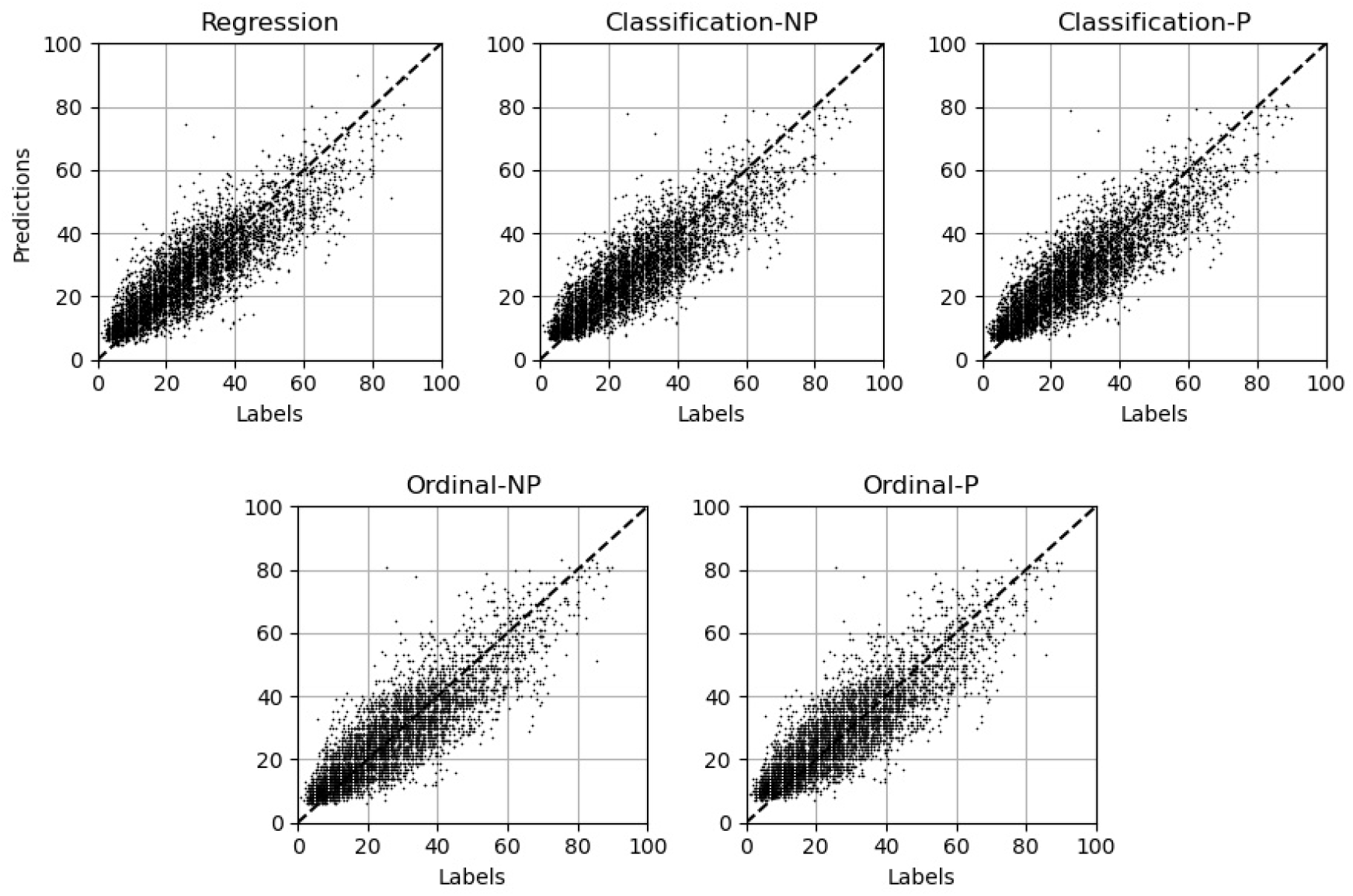
Predictions plotted against labels for the test set. Only 5,000 results out of a possible 25,158 are shown to make viewing easier.

**Figure A2.**
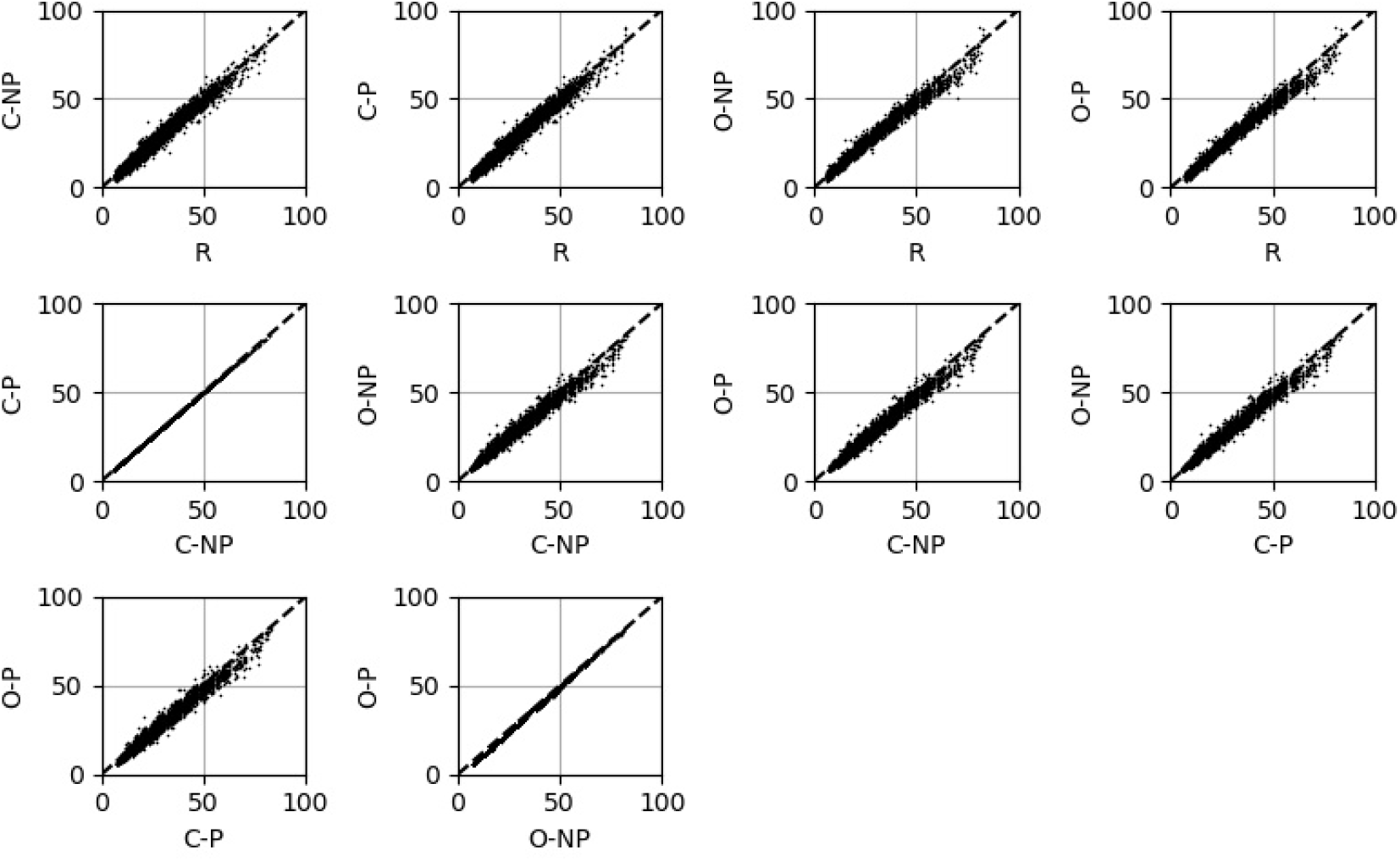
Model predictions plotted against one another to show the direct similarities and differences in model predictions.

**Figure A3.**
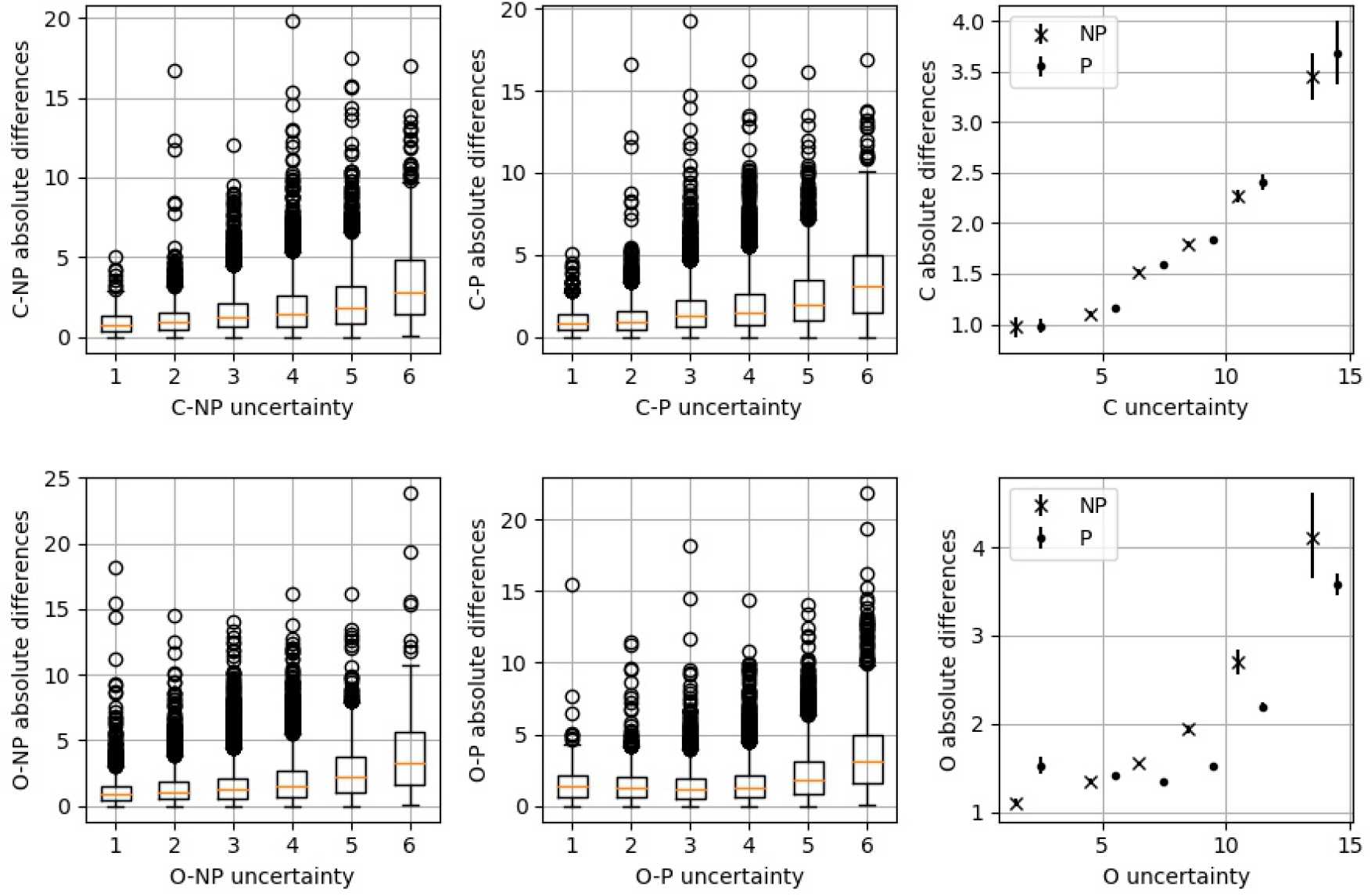
The model uncertainties are split into six bins and the absolute differences between the regression predictions and classification/ordinal predictions are shown. Left and middle) boxplots showing the absolute differences for the different uncertainty binned results. Right) averages of the absolute differences between classification/ordinal predictions and regression predictions in each of the uncertainty bins. The uncertainty bounds are via bootstrapping at the 95% level. Top plots are for the classification model and bottom plots for the ordinal model.

## Acknowledgements and funding

This work was funded by Canadian Institutes of Health Research (CIHR grant #169005). D. Gareth Evans and Susan M. Astley are supported by the National Institute for Health Research (NIHR) Manchester Biomedical Research Centre (Grant No. IS-BRC-1215-20007). Anne Martel is partially supported by the Tory Family Chair in Oncology. Steven Squires was affiliated with the University of Manchester whilst this work was done and is now affiliated with the University of Exeter.

## Code and data availability

Related code is available at https://github.com/stevensquires.

The PROCAS data is not available to researchers outside the University of Manchester.

